# Absorption Kinetics of Vitamins and Minerals from a Novel Nutritional Product in Healthy Adults: A Randomized, Double Blind, Placebo-Controlled Crossover Trial

**DOI:** 10.64898/2026.01.05.25342664

**Authors:** Philip A. Sapp, Jeremy R. Townsend, Trevor O. Kirby, Caitlyn G. Edwards, Michael B. La Monica, Tim N. Ziegenfuss, Matthew J. Vernge, Wendell S. Akers, Ralph Esposito

## Abstract

**Background/Objectives:** Nutrient interactions in multi-ingredient supplements may influence absorption and bioavailability, yet pharmacokinetic data in this context remains limited. This clinical trial assessed the post-prandial absorption kinetics of key micronutrients in AG1, a comprehensive supplement containing vitamins, minerals, probiotics, and phytonutrients.

**Methods:** In a randomized, double-blind, placebo-controlled crossover trial 16 healthy adults (8 males and 8 females) consumed a single serving (13g) of AG1 or a taste- and appearance-matched placebo mixed in water, following a 10-hour overnight fast. Each condition was separated by a 1-week washout. Blood samples were collected pre-consumption and at 30, 60, 90, 120, 180, 240, 360, and 480 minutes post-ingestion. Plasma concentrations of folate, calcium, zinc, vitamin C, biotin, nicotinamide, pyridoxine, riboflavin, thiamin, and hesperidin were measured. Area under the curve (AUC_0-480 min_) was used to assess nutrient absorption. Safety and tolerability were assessed throughout the study. Statistical analysis included repeated measures ANOVA and paired t-tests.

**Results:** AG1 significantly increased AUC_0-480 min_ values (p<0.05) for all measured nutrients except pyridoxine which revealed a strong trend (p = 0.075) and hesperidin (p = 0.224). Both AG1 and placebo were well tolerated, with no serious adverse events reported.

**Conclusions:** Acute consumption of AG1 resulted in measurable increases in circulating levels of most of the tested micronutrients, indicating effective absorption and bioavailability. These findings support the potential of AG1 to contribute meaningfully to nutritional status and overall health.

## 1. Introduction

Dietary supplements are widely consumed in the United States, with more than half of adults (61.4%) reporting regular dietary supplement use[1]. The most commonly consumed dietary supplement category is multivitamin–mineral supplements (MVMs), or supplements that contain three or more vitamins and one or more minerals [1]. Data from the National Health and Nutrition Examination Survey (NHANES) reveals that dietary supplements are often consumed by US adults with the intent to “improve” (45% of adults) or “maintain” overall health (33%), with MVMs specifically taken by many to “supplement the diet” [2,3]. Evidence consistently supports the ability of MVM supplements to help meet nutrient recommendations and improve indices of nutrient status[4–7].

Despite widespread MVM use and a demonstrated ability to improve dietary nutrient adequacy, there is a notable lack of data on the absorption kinetics and bioavailability of their constituent vitamins and minerals, both individually and in combination within the MVM matrix. The lack of these data were identified as a key area requiring further research by the 2010 Dietary Guidelines Advisory Committee [8].The committee called for the need to “develop accurate composition and bioavailability data across the multitude of vitamin, mineral, and nutrient supplements, and evaluate outcomes based on nutrient composition and bioavailability within the MVM matrix.” In response, a review by Comerford et al. [9] highlighted the challenges associated with each of these research areas identified in the 2010 DGAC report and the limitations of their referenced studies, suggesting the need for more clinical trials assessing the absorption kinetics of the vitamins and minerals in MVMs and evaluating the bioavailability of MVMs in different forms. However, the review emphasized the complexity and nuances of assessing the bioavailability or pharmacokinetics of MVMs compared to pharmaceuticals given their differences in composition and metabolism [9].

Despite the call for more research from the 2010 DGAC committee, published data on the absorption kinetics of multi-ingredient formulations remain limited. A small number of bioavailability studies assessing MVMs exist, [10–12], however, more recent additions to the literature focused on absorption kinetics continue to be scarce [13–16]. The complexity of the MVM matrix and inclusion of other bioactives in multi-ingredient products introduce multiple factors that may influence nutrient absorption. For example, nutrient-nutrient [17] and nutrient-matrix [18]interactions can modulate nutrient absorption and bioavailability, underscoring the need to examine how ingredient combinations and matrix composition influences in vivo absorption.

The multi-ingredient dietary supplement, AG1, is a daily foundational nutritional supplement that combines vitamins, minerals, phytonutrients, prebiotics, and probiotics, with whole food powders and other similar ingredients. Within complex matrices like these, it is unknown if certain vitamins and minerals exhibit delayed, impaired, or improved absorption due to physical and chemical interactions, prompting the need to characterize the absorption kinetics of key nutrients in full formulations. A prior formulation of AG1 was assessed using the Simulator of the Human Intestinal Microbial Ecosystem® (SHIME), which demonstrated that its minerals were bioaccessible and likely to enter systemic circulation in vitro. However, the model’s constraints limited accurate assessment of a wider array of nutrients which are more appropriately measured in vivo [19]. Thus, validating the plasma appearance and kinetic parameters of AG1’s nutrients is essential to substantiate its supplemental efficacy.

Therefore, the present study was designed to quantitatively assess the plasma appearance and absorption kinetics of key nutrients in AG1. We conducted a double-blind, randomized, placebo-controlled crossover trial in healthy men and women to measure the plasma appearance of several vitamins and minerals from AG1. Primary outcomes included folate, calcium, zinc, and vitamin C. Secondary outcomes consisted of B-vitamins, dietary nutrient adequacy, tolerability, and safety. By characterizing the plasma appearance of these nutrients over an 8-hour period post-consumption, we aimed to evaluate some of the absorption kinetics of AG1 and contribute to the growing body of literature on multi-ingredient supplement pharmacokinetics. This study addresses a gap in pharmacokinetic data of complex formulations and evaluates whether AG1 can effectively deliver bioavailable nutrients via its MVM matrix.

## 2. Materials and Methods

### 2.1. Participants

Sixteen healthy men (n = 8) and women (n = 8) completed this trial. Eligible participants were enrolled if they were in good health based on their health history, physical examination, and blood chemistries. Participants were ages 18-45 years, had a body mass index of 18.5-29.9 kg/m2, had normotensive blood pressure (systolic ≤140 mm Hg and diastolic ≤ 90 mm Hg), had a seated resting heart rate ≤ 90 beats per minutes, and were willing to duplicate their baseline dietary intake, refrain from caffeine and exercise for 24 hours prior, and fast for 10 hours prior to each testing visit. Before enrollment, all participants indicated their willingness to comply with all aspects of experimental protocol. Exclusion criteria included: (a) MVM supplementation within the past 3 months or any other dietary supplement which would interfere with study outcomes (e.g, vitamins, minerals, probiotics, prebiotics, etc.) (b) subjects who have received an antibiotic within 3 months prior to study entry or current use of prescription or OTC medications that could influence study outcomes; (c) presence of chronic disease or medical condition such as cancer, gastrointestinal, cardiovascular, respiratory, liver, renal, thyroid, autoimmune, or metabolic conditions; (d) alcohol consumption (more than 2 standard alcoholic drinks per day or more than 10 drinks per week), drug abuse or dependence within the past 6 months; (e) current smokers, vapers, or tobacco users or use of tobacco within the past month (f) pregnant women, women trying to become pregnant, women less than 120 days postpartum or nursing women. (g) a change in hormone therapy, including oral contraceptives, within 4 weeks prior to screening, or unwilling to maintain current hormone therapy/oral contraceptive use throughout the course of the study; (h) known sensitivity to any ingredient in the test formulations as listed in the product label. Participants were also excluded if they were currently participating in another research study with an investigational product or have been in another research study in the past 30 days, adhered to subject is a specialty diet including but not limited to, Vegan, Vegetarian, Ketogenic, Paleo, Atkins, South Beach, Carnivore, etc; or had donated blood or plasma within the previous week.

This study was conducted according to the guidelines in the Declaration of Helsinki of 1975, and all study procedures were approved by the WCG Institutional Review Board (#AG-02-0124). Prior to enrollment, written informed consent was obtained from all study participants.

### 2.2. Experimental Design

This was a double-blind, randomized, placebo-controlled crossover clinical trial. Participants attended three laboratory visits throughout the study: a screening visit and two testing visits to complete the pharmacokinetic assessments on AG1 and placebo (PL) (Figure 1). AG1 and PL conditions were separated by a one-week, or greater, washout period. Participants were randomized to their respective treatment sequence. Both participants and investigators were blinded to the study products. This study was registered on clinicaltrials.gov (NCT06316700).

**Figure 1.**
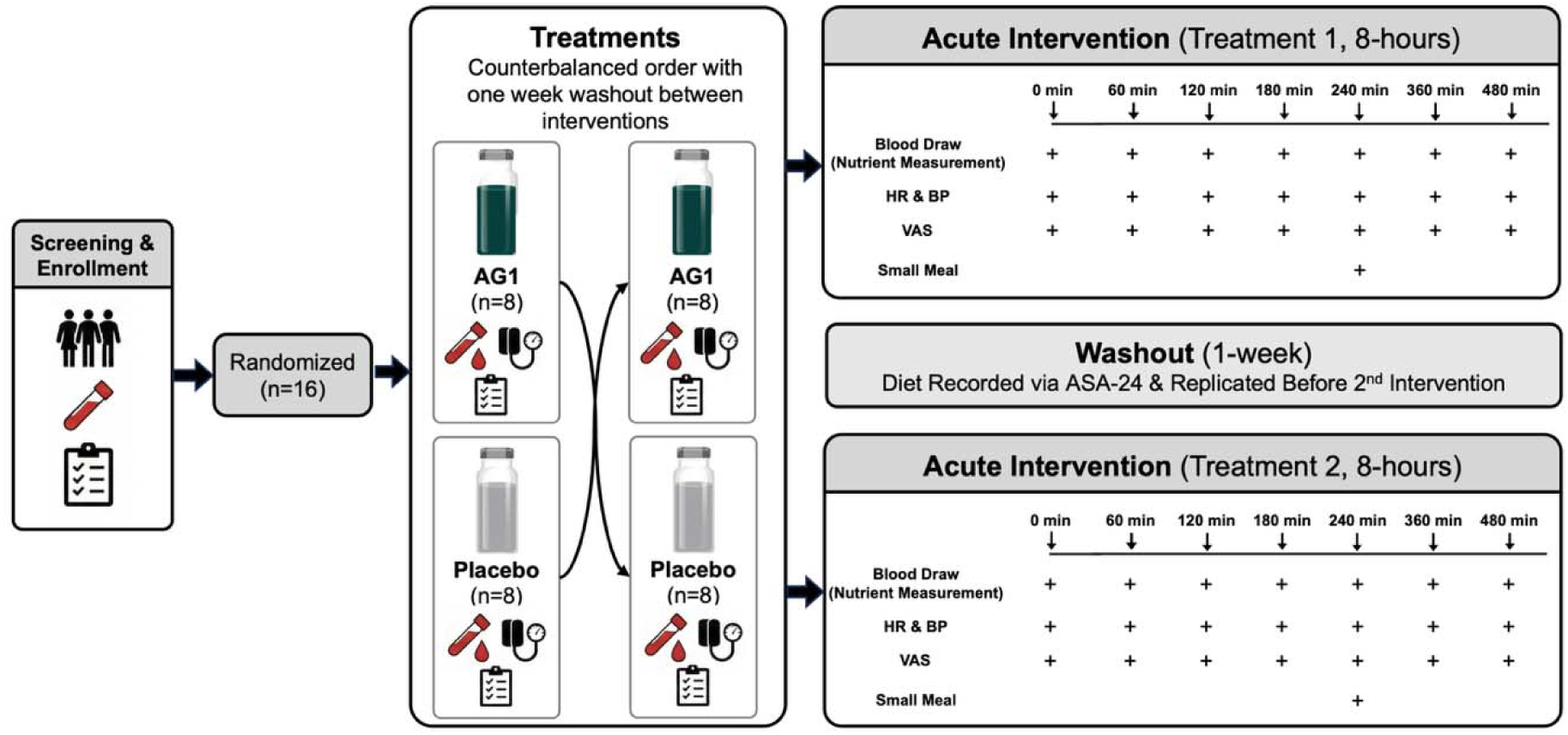
Study diagram detailing the study flow for subjects participating in this study. BP, blood pressure; HR, heart rate; min, minutes; and VAS, visual analog scale.

### 2.5. Blood Sample Collection and Handling

Blood draws took place during each study visit. At screening a single blood draw was collected with a total blood volume of approximately 12 mL. During visits 2 and 3 serial blood draws were completed using an intra-venous catheter was inserted into an upper extremity vein before and 30-, 60-, 90-, 120-, 180-, 240-, 360-, and 480 min post ingestion of the investigational product. The blood handling procedures were as follows at each timepoint: 1 serum separator tube (allowed to clot for 30 minutes and spun in LabCorp centrifuge then sent to LabCorp for calcium and folate analysis), 1 large heparin coated tube (immediately spun twice in LabCorp centrifuge, split into 4 separate 0.5 mL aliquots, stored at −80 degrees Celsius), 1 small heparin coated tube (immediately spun in LabCorp centrifuge, aliquoted 2 mL into dark cryovial, stored for >24 hours at −80 degrees Celsius, and sent to LabCorp for vitamin C analysis), and 1 trace element K2 EDTA tube (im-mediately spun in Eppendorf for 15 minutes at 3.0 RPM and 20 degrees Celsius, aliquoted 2 mL to metal free transfer tube, and sent to LabCorp for zinc analysis). Additional aliquots for analysis of the remaining blood markers were stored at −80 degrees Celsius and were analyzed after the completion of the study.

### 2.6. Laboratory Analysis of Analytes

A subset of the prepared blood specimens were submitted to LabCorp (Dublin, OH, USA) for the lipid panel (Test Number 303756), CBC with differential (Test Number 005009), comprehensive metabolic panel (Test Number 322000), total serum folate(Test Number 002014), total serum zinc (Test Number 001800), total plasma vitamin C (Test Number 001805), and total serum calcium (Test Number 001016) analysis. The lipid panel included total cholesterol, HDL cholesterol, LDL cholesterol, triglycerides, and VLDL cholesterol. The CBC with differential panel included hematocrit, hemoglobin, mean corpuscular volume (MCV), mean corpuscular hemoglobin (MCH), mean corpuscular hemoglobin concentration (MCHC), red cell distribution width (RDW), percentage and absolute differential counts, platelet count (PLT), red cell count (RBC), and white blood cell count (WBC). The comprehensive metabolic panel included alanine aminotransferase (ALT/SGPT), albumin:globulin (A:G) ratio, serum albumin, serum alkaline phosphatase, aspartate aminotransferase (AST/SGOT), total bilirubin, BUN, BUN:creatinine ratio, serum calcium, total carbon dioxide, serum chloride, serum creatinine, eGFR calculation, total globulin, serum glucose, serum potassium, total serum protein, and serum sodium. All lab analyses were conducted using LabCorp’s standard operating procedures, with more information available through their website.

The remaining blood specimens were submitted to Lipscomb University (Nashville, TN, USA) to measure serum levels of a vitamin B panel, serum phytochemicals, and total antioxidant capacity. The serum vitamin B panel consisted of thiamine, biotin, riboflavin, nicotinamide, methylcobalamin, and pyridoxine. The phytochemical panel consisted of hesperidin, a citrus bioflavonoids. Calibration and QC standards were prepared in concentrations of 0.1 ng/mL to 1000 ng/mL for the mixture of each vitamin and citrus bioflavonoid. Internal standard calibration curves were prepared for each analyte.

A simple protein precipitation procedure was employed for sample preparation. To 100 μL of plasma sample (calibration standards, QC samples, or study samples), 300 μL of internal standard solution (100 ng/mL internal standard mix in acetonitrile) was added in a well on a 96-well protein precipitation plate (Restek Resprep PPT3, Bellefonte, PA). The mixture was vortexed for 30 seconds, and positive pressure was used to collect filtrate into a 96-well deep well sample plate. The filtrate was dried with a stream of nitrogen, re-constituted with 100 μL of water, and vortexed briefly before being transferred to the LC-MS/MS for analysis.

Chromatographic separation was performed on a Shimadzu Prominence HPLC system (Shimadzu, Columbia, MD) equipped with a binary pump, autosampler, and column oven. Mass spectrometric detection was carried out using a Shimadzu LCMS 8040 triple quadrupole mass spectrometer equipped with a Dual Electrospray/Atmospheric Pressure Chemical Ionization (DuIS). Chromatographic separation was achieved on a Restek Aqua Ultra Aqueous C18 column (50 × 2.1 mm, 3 μm) maintained at 40°C. The mobile phase consisted of (A) 0.1% formic acid and 5 mM ammonium formate in water and (B) 0.1% formic acid in 5 mM ammonium formate in acetonitrile. The flow rate was 0.6 mL/min with a total run time of 6 minutes. The gradient program was as follows: 0.0-2.0 min, 0-65% B; 2.01 min, 90% B; 2.01-4.0 min, 90-95% B; 4.0-4.5 min, 95% B; 4.51 min, 0% B (re-equilibration). The injection volume was 20 μL.

The mass spectrometer was operated in positive ionization (+) mode for all analytes. The optimized source parameters were: Desolvation Line 250 °C, Heat Block 400°C, and Drying Gas Flow 20 L/min, and Nebulizing gas flow 3 L/min. Compound-specific parameters, including precursor ion, product ion, Q1 pre-rod bias, collision energy (CE), and Q3 pre-rod bias, were optimized for each analyte and internal standard (Table S1). The concentrations were determined by comparing the sample peak area ratio to the corresponding calibration curve for each analyte.

### 2.7. Subjective Outcome Assessments

Participants were asked to complete Visual Analog Scales (VAS) during visit two and three at baseline, 30-, 60-, 90-, and 120-min post-ingestion of the investigational product. These were used to assess levels of bloating, hunger, flatulence (gas), fullness (satiety), energy, sleepiness, desire to eat (appetite), prospective food consumption, and overall mood. A standardized 10 cm line was used with the left end of each line anchored with descriptors depicting the lowest possible level/score and the highest possible level/score descriptors were anchored to the right of each line. Following data collection, the VAS responses were quantified by measuring the distance from the zero anchor to each participant’s mark in millimeters (range: 0-10 cm for each metric).

### 2.8. Pharmacokinetic Calculations

The key pharmacokinetic parameters measured included maximum plasma con-centration observed (C_max_), time to maximum plasma concentration was observed (T_max_), and total plasma exposure for up to 8 hours (AUC_0-480 min_). Maximal concentration was determined by using the highest observed concentration, with the time to maximum concentration simply being the time point in which Cmax was observed. Total plasma exposure for up to 8 hours was calculated via Area Under the Curve (AUC) utilizing the trapezoid rule (GraphPad Software version 10.6.0 for Windows, Boston, Massachusetts USA). For the analysis, the baseline concentration of a respective nutrient was used in AUC calculations. Total AUC was utilized to determine the total plasma exposure of a given nutrient.

### 2.9. Safety Monitoring and Adverse Events

All local and systemic non-serious and serious adverse events (AEs) were reported by the investigators assessed through reports coded using the Medical Dictionary for Regulatory Activities (MedDRA). The intensity of an AE was subsequently graded according to the protocol-defined criteria based on the Common Terminology Criteria for Adverse Events (CTCAE) Version 5.0, 2017.

### 2.10. Statistical Analyses

Based on previous pharmacokinetic studies examining these nutrients of interest, a minimum sample size of N=10 was chosen to detect statistically significant changes between treatments with differences in AUC0-t as the primary outcome variable. Specifically, assuming an alpha of 0.05, beta of 0.80, and effect size of 1.78 [based on the nutrient with the smallest effect size], we calculated a sample size estimate of 5 participants per sex. Accounting for dropouts and participant variability we aim to enroll 8 men and 8 women. Secondary outcome variables included Cmax, Tmax, and the individual differences in the nutrient’s concentration at each sampling point. Additionally, tolerability and safety measures were assessed for both conditions.

Descriptive statistics (mean and SD) were used to quantify subjects’ baseline physical characteristics as well as their clinical chemistry and hematological biomarkers. Urine specific gravity, body weight, and dietary intake at baseline for both conditions were assessed with paired T-tests. Paired T-test was utilized to analyze the primary outcome (AUC_0-480 min_) as well as the kinetic parameters Cmax and Tmax comparing AG1 and the placebo group. Two-way ANOVA was utilized to compare the concentrations of each nutrient at each time point between AG1 and the placebo group. Data on VAS results are presented as mean ± SD and were assessed using two-way ANOVA to compare participant reported scores at each time point between AG1 and the placebo group. Similarly, blood pressure and heart rate results are presented as mean ± SD and were assessed using two-way ANOVA to compare the measures at each time point between both conditions. Sidak post hoc comparisons were made upon statistical significance in the mixed factorial ANOVA model. All variables were tested for normality using results from a Shapiro-Wilk test. For all statistical tests, significance was set at P < 0.05. All statistical analyses were conducted using GraphPad Prism version 10.1.1.

## 3. Results

### 3.1. Participants

All 16 enrolled participants completed the study. Participants consisted of 8 healthy males (27.4 ± 8.9 yrs; 81.5 ± 11.8 kg) and 8 healthy females (33.1 ± 9.7 yrs; 68.9 ± 6.0 kg). Baseline characteristics, anthropometrics and blood chemistry values are presented in Tables 1 and Table *S2*.

**Table 1.** Baseline demographics and clinical characteristics of the study participants.

| Variable | Total (n = 16) | Males (n = 8) | Females (n = 8) |
| --- | --- | --- | --- |
| Age (years) | 30.3 ± 9.5 | 27.4 ± 8.9 | 33.1 ± 9.7 |
| Height (cm) | 171.4 ± 10.0 | 178.8 ± 7.8 | 164.0 ± 5.5 |
| Weight (kg) | 75.2 ± 11.1 | 81.5 ± 11.8 | 68.9 ± 6.0 |
| Body Mass Index (kg/m<sup>2</sup>) | 25.2 ± 2.0 | 24.7 ± 1.9 | 25.8 ± 2.1 |
| Systolic Blood Pressure (mm Hg) | 116.4 ± 13.7 | 121.1 ± 14.0 | 111.6 ± 12.3 |
| Diastolic Blood Pressure (mm Hg) | 71.3 ± 6.9 | 68.4 ± 5.5 | 74.1 ± 7.3 |
| Resting Heart Rate (bpm) | 67.6 ± 14.2 | 62.3 ± 13.0 | 72.9 ± 14.0 |

Values are presented as mean ± SD for the total sample (n = 16). Bpm, beats per minute; Cm, centimeters; kg, kilograms; kg/m<sup>2</sup>, kilograms per meter<sup>2</sup>; and mm Hg, millimeters of mercury.

### 3.2. Vitamin and Mineral Results

#### 3.2.1 Folate

Folate (Figure 2 Panels A and B and Table S3) concentrations were significantly elevated at 30, 60, 90, 120, 180, 240, 360, and 480 minutes post ingestion for AG1 relative to placebo. When AG1 was ingested AUC_0-480min_, C_max_, and T_max_ were 8,124 ±1,297 ng/mL/min, 20.8 ± 0.7 ng/mL, and 37.5 ± 17.3 min respectively. When the placebo was ingested AUC_0-480min_, C_max_, and T_max_ were 5,489 ± 1,723 ng/mL/min, 12.9 ± 3.9 ng/mL, and 174 ± 111 min respectively. All three kinetic parameters were significantly different between the two treatment groups (each exhibiting P < 0.001).

**Figure 2.**
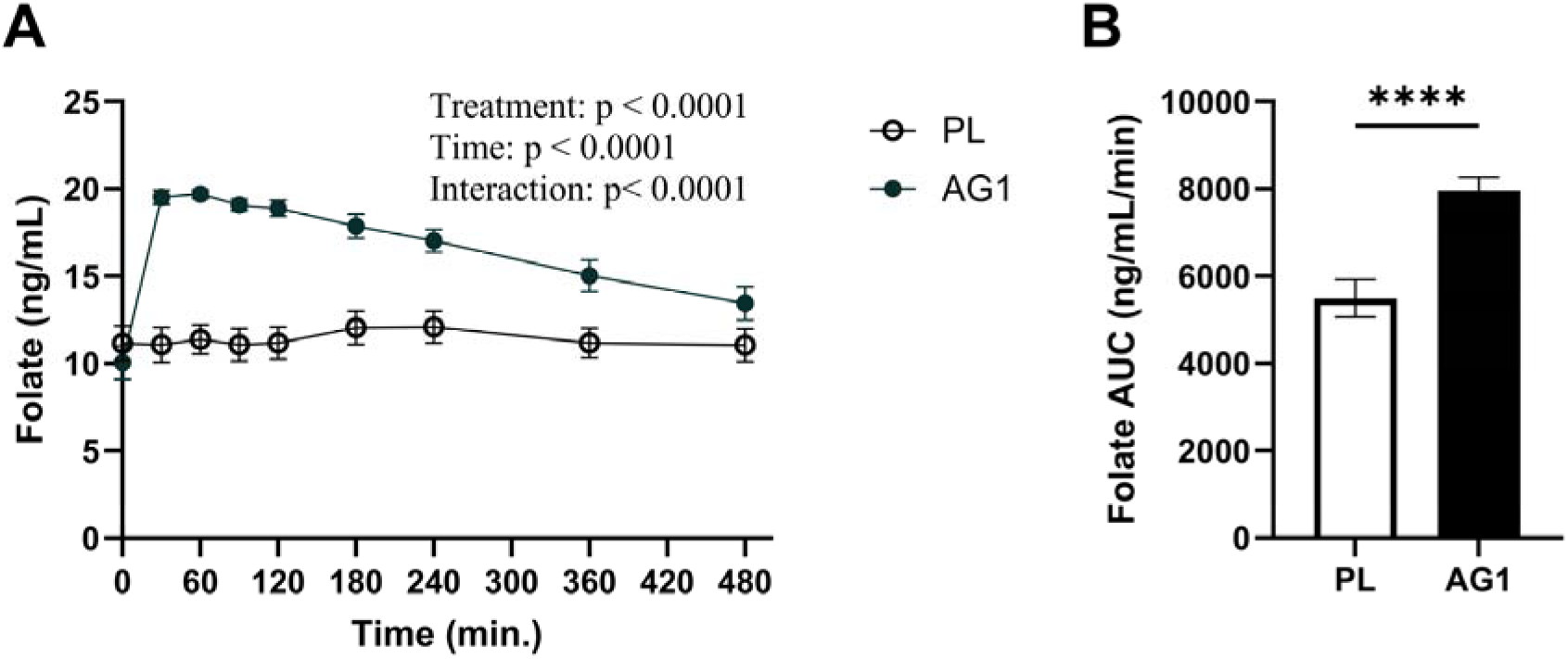
Folate concentrations and total AUC. Visualization of changes in the folate curves over the duration of 480 minutes (A) and total AUC (B). ng/mL, nanogram/milliliter; ng/mL/min, nanogram/milliliter/minute; and PL, placebo. ****, p < 0.0001.

#### 3.2.2. Calcium

Calcium (Figure 3 Panels A and B and Table S3) concentrations were significantly elevated at 30, 60, 90, 120, 180, and 240 minutes post ingestion for AG1 relative to placebo. When AG1 was ingested AUC_0-480min_, C_max_, and T_max_ were 4,448 ± 116 mg/dL/min, 9.51 ± 0.26 mg/dL, and 129 ± 88.8 min respectively. When the placebo was ingested AUC_0-480min_, C_max_, and T_max_ were 4,366 ± 146 mg/dL/min, 9.38 ± 0.3 mg/dL, and 229 ± 167 min respectively. With respect to the kinetic parameters, AUC_0-480min_ is significantly different between the two treatment groups (P < 0.01) as well as C_max_ and T_max_ (P < 0.05).

**Figure 3.**
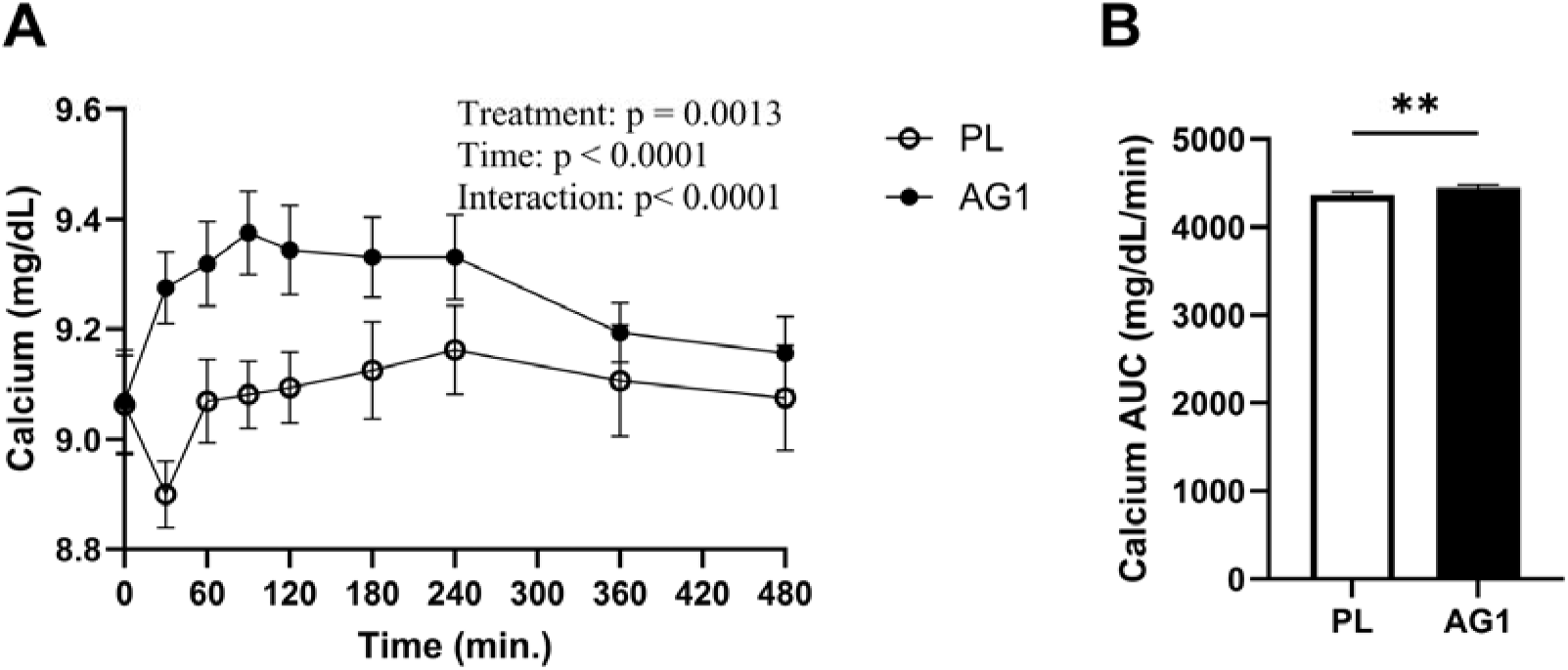
Calcium concentrations and total AUC. Visualization of changes in the calcium curves over the duration of 480 minutes (A) and total AUC (B). mg/dL, milligram/deciliter; mg/dL/min, milligram/deciliter /minute; and PL, placebo. **, p < 0.01.

#### 3.2.3. Zinc

Zinc (Figure 4 Panels A and B and Table S3) concentrations were significantly elevated at 60, 90, 120, 180, and 240 minutes post ingestion for AG1 relative to placebo. When AG1 was ingested AUC_0-480min_, C_max_, and T_max_ were 41,140 ± 2,938 μg/dL/min, 112 ± 14 μg/dL, and 150 ± 68 min respectively. When the placebo was ingested AUC_0-480min_, C_max_, and T_max_ were 35,688 ± 3,353 μg/dL/min, 87.1 ± 8.8 μg/dL, and 104 ± 88.5 min respectively. With respect to the kinetic parameters, AUC_0-480min_ and C_max_ were significantly different between the two treatment groups (P < 0.001), but no significance was observed for T_max_ (P > 0.05).

**Figure 4.**
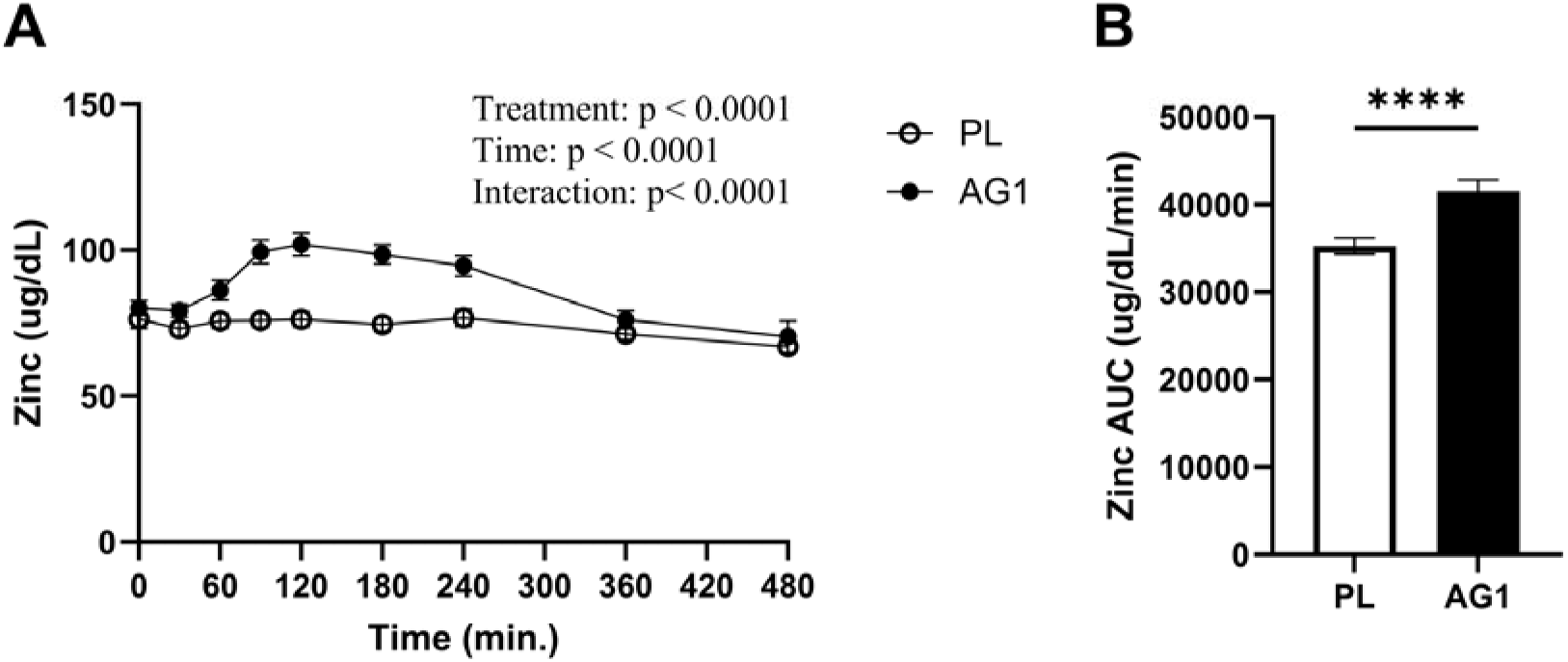
Zinc concentrations and total AUC. Visualization of changes in the zinc curves over the duration of 480 minutes (A) and total AUC (B). ug/dL, microgram/deciliter; ug/dL/min, microgram/deciliter /minute; and PL, placebo. ****, p < 0.0001.

#### 3.2.4. Vitamin C

Vitamin C (Figure 5 Panels A and B and Table S3) concentrations were significantly elevated at 30, 60, 90, 120, 180, 240, 360, and 480 minutes post ingestion for AG1 relative to placebo. When AG1 was ingested AUC0-480min, Cmax, and Tmax were 598 ± 276 mg/dL/min, 1.55 ± 0.71 mg/dL, and 146 ± 96 min respectively. When the placebo was ingested AUC0-480min, Cmax, and Tmax were 358 ± 206 mg/dL/min, 0.93 ± 0.49 mg/dL, and 135 ± 144 min respectively. With respect to the kinetic parameters, AUC0-480min and Cmax were significantly different between the two treatment groups (P < 0.001) but no significance was observed for Tmax (P > 0.05).

**Figure 5.**
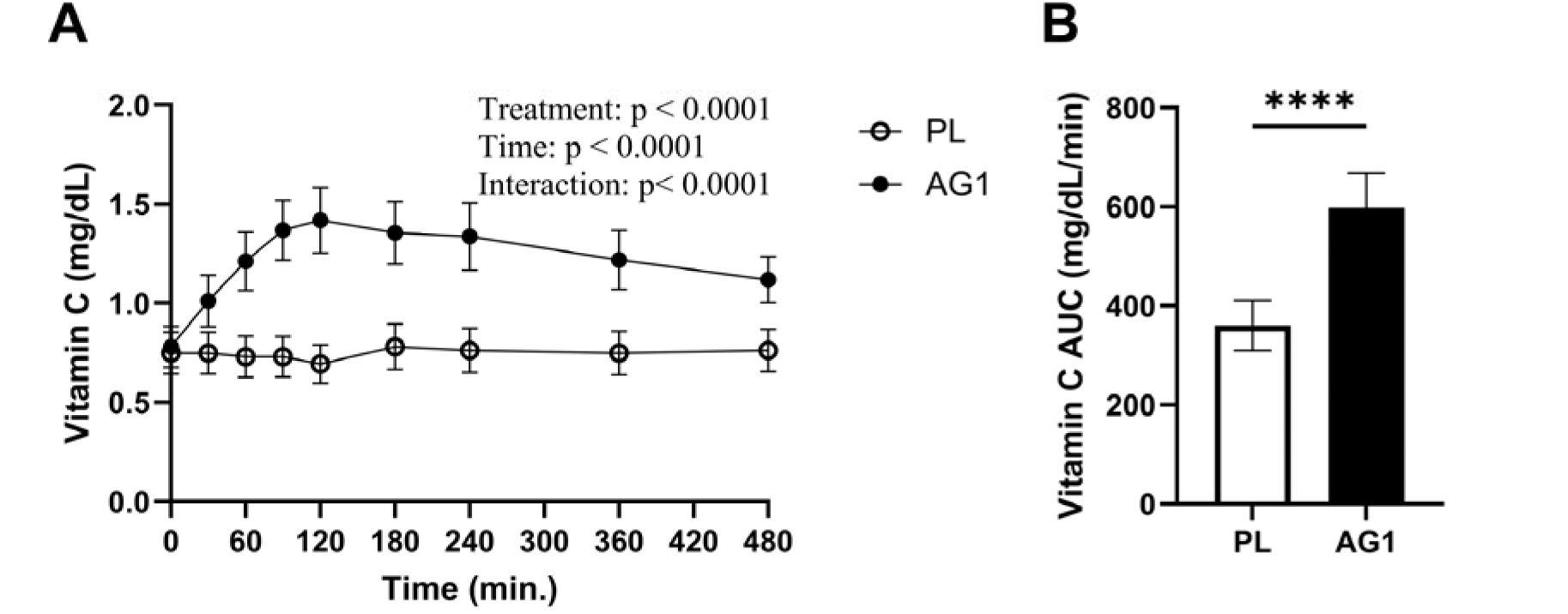
Vitamin C concentrations and total AUC. Visualization of changes in the vitamin C curves over the duration of 480 minutes (A) and total AUC (B). mg/dL, milligram/deciliter; mg/dL/min, milligram/deciliter /minute; and PL, placebo. **, p < 0.01.

#### 3.2.5. Biotin

Biotin (Figure 6 Panels A and B and Table S3) concentrations were significantly elevated at 30, 60, 90, 120, and 180 minutes post ingestion for AG1 relative to placebo. When AG1 was ingested AUC_0-480min_, C_max_, and T_max_ were 404.4 ± 298.3 mg/dL/min, 2.26 ± 1.9 mg/dL, and 58 ± 32 min respectively. When the placebo was ingested AUC_0-480min_, C_max_, and T_max_ were 154 ± 225.1 mg/dL/min, 0.57 ± 0.68 mg/dL, and 139 ± 181 min respectively. With respect to the kinetic parameters, AUC_0-480min_ and C_max_ were significantly different between the two treatment groups (P < 0.0001 and P = 0.0036, respectively) but no significance was observed for T_max_ (P > 0.05).

**Figure 6.**
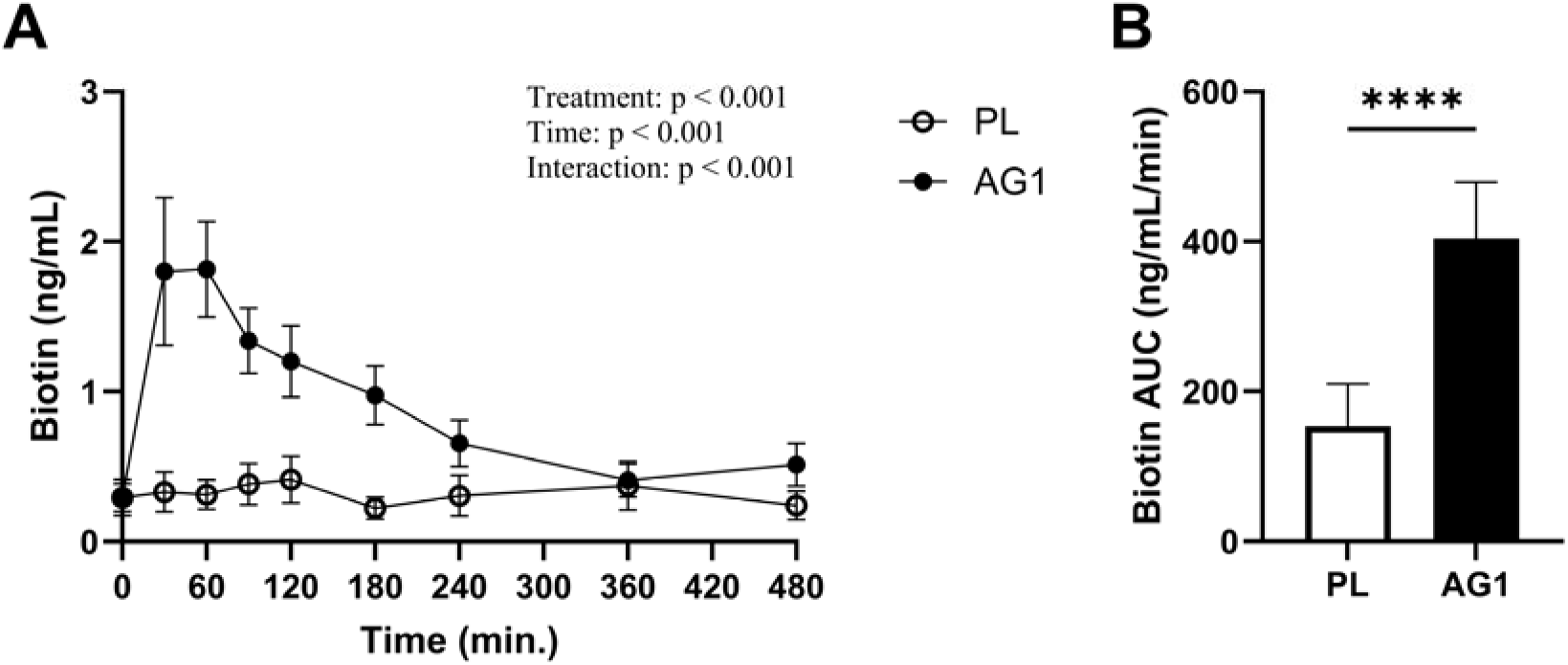
Biotin concentrations and total AUC. Visualization of changes in the biotin curves over the duration of 480 minutes (A) and total AUC (B). ng/mL, nanogram/milliliter; ng/mL/min, nanogram/milliliter/minute; and PL, placebo. ****, p < 0.0001.

#### 3.2.6. Riboflavin

Riboflavin (Figure 7 Panels A and B and Table S3) concentrations were significantly elevated at 30 minutes post ingestion for AG1 relative to placebo. When AG1 was ingested AUC_0-480min_, C_max_, and T_max_ were 3,271.4 ± 6,319.3 ng/mL/min, 19.6 ± 25.7 ng/mL, and 79 ± 83 min respectively. When the placebo was ingested AUC_0-480min_, C_max_, and T_max_ were 1,434.9 ± 5,058.7 ng/mL/min, 15.2 ± 40.9 ng/mL, and 206 ± 143 min respectively. With respect to the kinetic parameters,AUC_0-480min_ (P < 0.001) and T_max_ (P < 0.05) were significantly different, while C_max_ was not significantly different (P > 0.05) between the two treatment groups.

**Figure 7.**
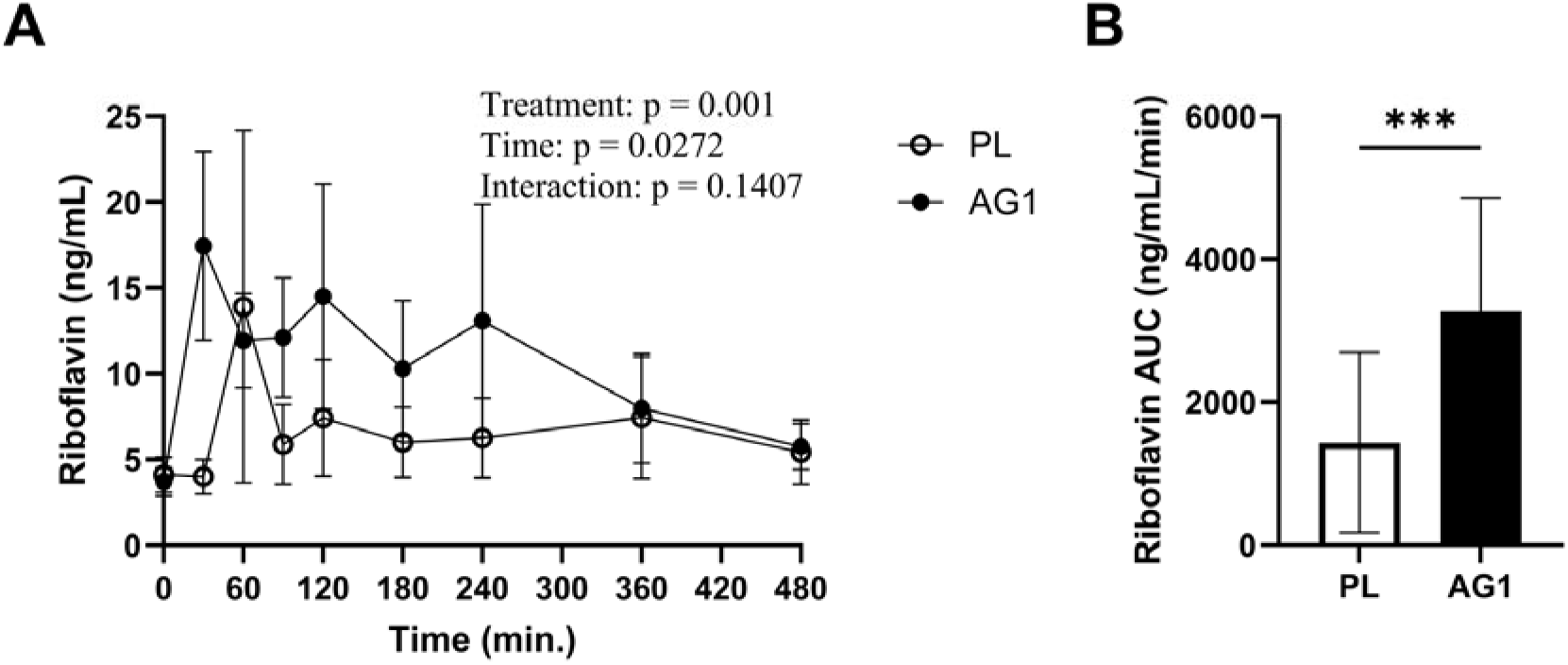
Riboflavin concentrations and total AUC. Visualization of changes in the riboflavin curves over the duration of 480 minutes (A) and total AUC (B). ng/mL, nanogram/milliliter; ng/mL/min, nanogram/milliliter/minute; and PL, placebo. ***, p < 0.001.

#### 3.2.7. Thiamine

Thiamine (Figure 8 Panels A and B and Table S3) concentrations were significantly elevated at 30, 60, 90, 120, 180, 240, and 360 minutes post ingestion for AG1 relative to placebo. When AG1 was ingested AUC_0-480min_, C_max_, and T_max_ were 3,047.9 ± 2,865.4 ng/mL/min, 17.35 ± 13.03 ng/mL, and 84 ± 49 min respectively. When the placebo was ingested AUC_0-480min_, C_max_, and T_max_ were 47.9 ± 116.8 ng/mL/min, 2.32 ± 2.49 ng/mL, and 174 ± 160 min respectively. With respect to the kinetic parameters, AUC_0-480min_ (P < 0.001), C_max_ (P < 0.0001) and T_max_ (P < 0.05) were significantly different between the two treatment groups.

**Figure 8.**
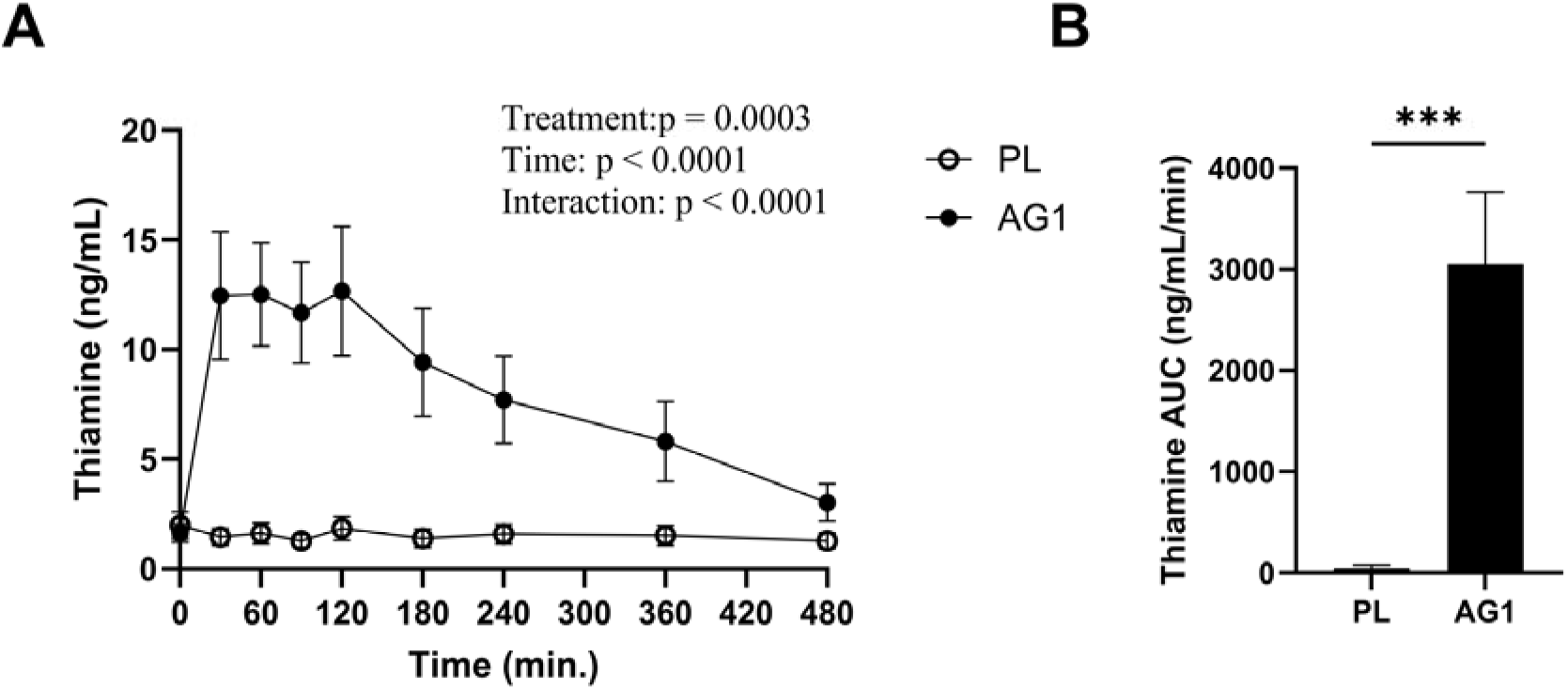
Thiamine concentrations and total AUC. Visualization of changes in the thiamine curves over the duration of 480 minutes (A) and total AUC (B). ng/mL, nanogram/milliliter; ng/mL/min, nanogram/milliliter/minute; and PL, placebo. ***, p < 0.001.

#### 3.2.8. Pyridoxine

Pyridoxine (Figure 9 Panels A and B and Table S3) concentrations were significantly elevated at 30 minutes post ingestion for AG1 relative to placebo. When AG1 was ingested AUC_0-480min_, C_max_, and T_max_ were 4.32 ± 4.1 ng/mL/min, 0.1 ± 0.07 ng/mL, and 58 ± 84 min respectively. When the placebo was ingested AUC_0-480min_, C_max_, and T_max_ were 2.19 ± 4.3 ng/mL/min, 0.07 ± 0.09 ng/mL, and 66 ± 69 min respectively. There was no significant difference observed for any of the kinetic parameters for AG1 or the placebo.

**Figure 9.**
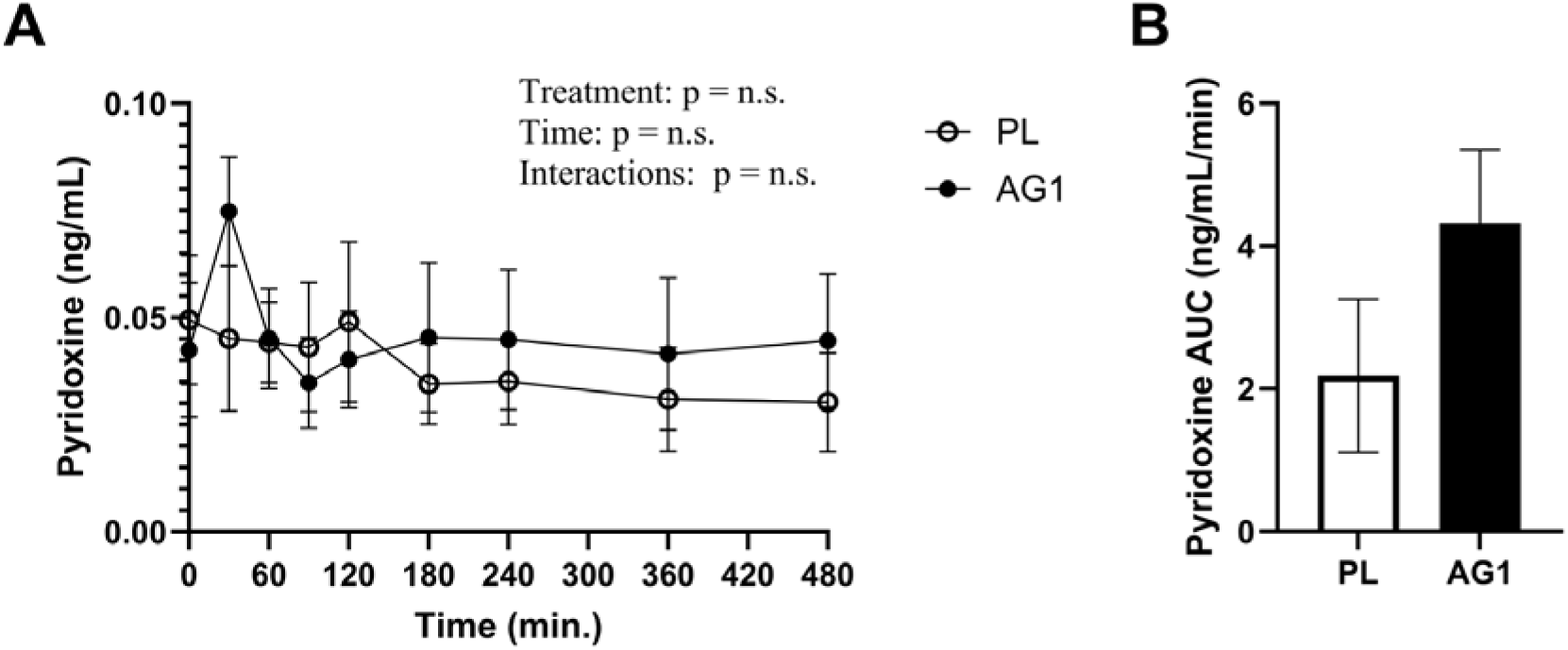
Pyridoxine concentrations and total AUC. Visualization of changes in the pyridoxine curves over the duration of 480 minutes (A) and total AUC (B). ng/mL, nanogram/milliliter; ng/mL/min, nanogram/milliliter/minute; and PL, placebo.

#### 3.2.9. Nicotinamide

Nicotinamide (Figure 10 Panels A and B and Table S3) concentrations were significantly elevated at 30, 60, 90, and 120 minutes post ingestion for AG1 relative to placebo. When AG1 was ingested AUC_0-480min_, C_max_, and T_max_ was 6,308.4 ± 7341.9 ng/mL/min, 81.6 ± 52.5 ng/mL, and 53 ± 57 min respectively. When the placebo was ingested AUC_0-480min_, C_max_, and T_max_ was 3,613.4 ± 6157.2 ng/mL/min, 38.77 ± 39.77 ng/mL, and 116 ± 65 min respectively. With respect to the kinetic parameters, AUC_0-480min_ (P < 0.05), C_max_ (P < 0.01) and T_max_ (P < 0.05) were significantly different between the two treatment groups.

**Figure 10.**
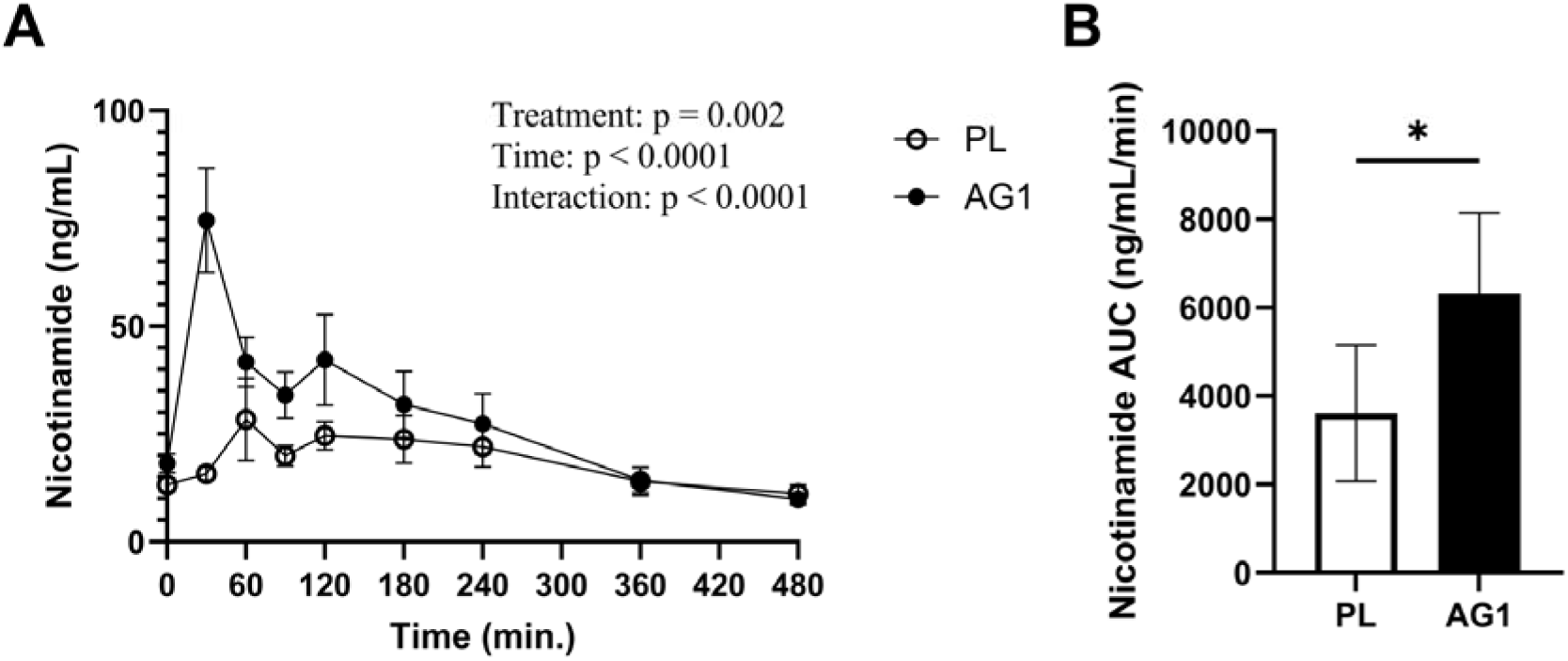
Nicotinamide concentrations and total AUC. Visualization of changes in the nicotinamide curves over the duration of 480 minutes (A) and total AUC (B). ng/mL, nanogram/milliliter; ng/mL/min, nanogram/milliliter/minute; and PL, placebo. *, p < 0.05.

#### 3.2.10. Hesperidin

Hesperidin (Figure 11 Panels A and B and Table S3) concentration was significantly elevated at 180 minutes post ingestion for AG1 relative to placebo. When AG1 was ingested AUC_0-480min_, C_max_, and T_max_ were 27.99 ± 70 ng/mL/min, 0.28 ± 0.75 ng/mL, and 129 ± 144 min respectively. When the placebo was ingested AUC_0-480min_, C_max_, and T_max_ were 5.32 ± 10.65 ng/mL/min, 0.07 ± 0.7 ng/mL, and 122 ± 151 min respectively. There was no significant difference observed for any of the kinetic parameters for AG1 or the placebo.

**Figure 11.**
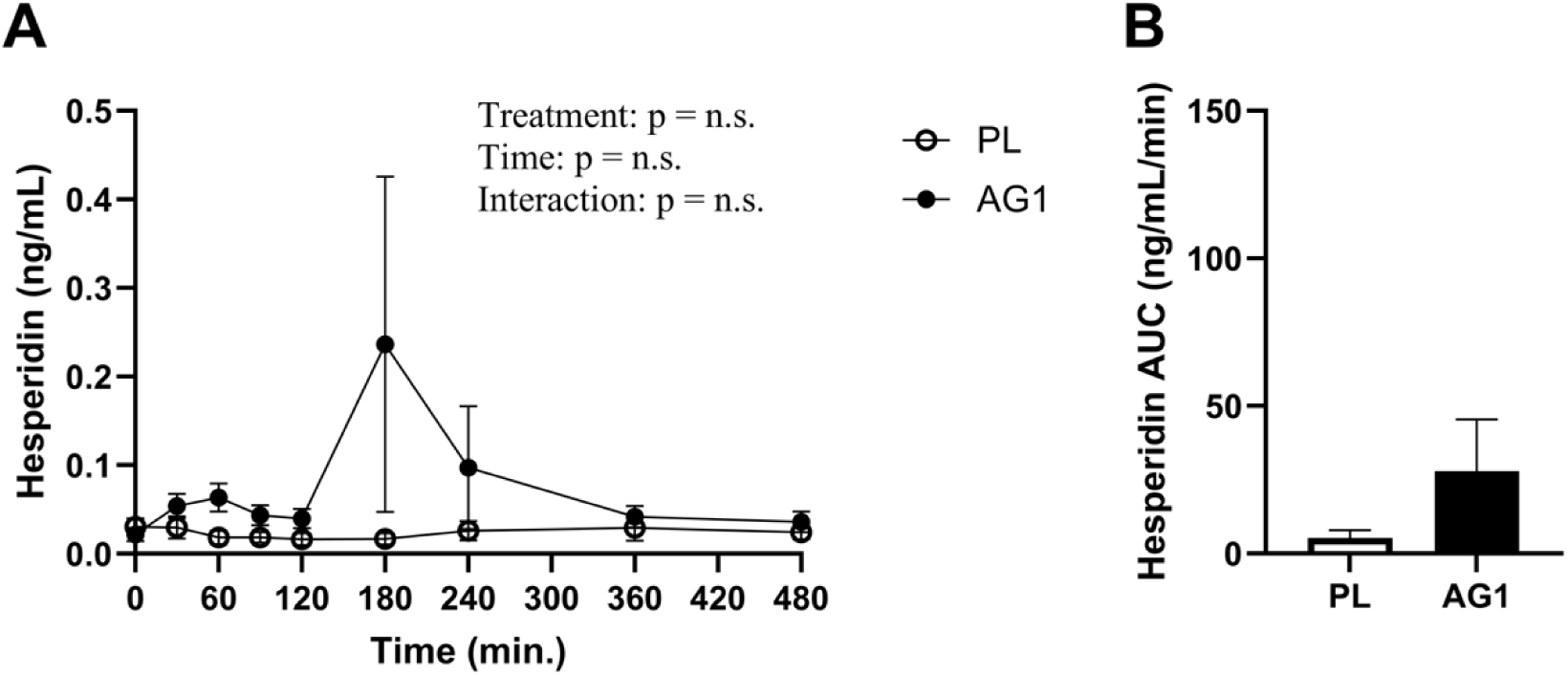
Hesperidin concentrations and total AUC. Visualization of changes in the hesperidin curves over the duration of 480 minutes (A) and total AUC (B). ng/mL, nanogram/milliliter; ng/mL/min, nanogram/milliliter/minute; and PL, placebo.

### 3.3. Tolerability and Safety

#### 3.3.1 Tolerability

No significant group x time interactions were observed for any of the VAS items (Table 2). A significant main effect for time and group was observed for hunger ratings (p<0.05). Hunger ratings were higher for the placebo group at 0 and 30 minutes when compared to AG1 (p=0.005 and p=0.015, respectively).

**Table 2.** Visual analog scale (VAS) ratings following consumption of each intervention.

| Variable | Group | 0 min | 30 min | 60 min | 90 min | 120 min |
| --- | --- | --- | --- | --- | --- | --- |
| Bloating (cm) | AG1 | 1.2 ± 1.2 | 1.3 ± 1.6 | 1.4 ± 1.5 | 1.1 ± 1.2 | 1.0 ± 1.0 |
|  | PL | 1.0 ± 1.2 | 1.5 ± 1.9 | 1.6 ± 2.0 | 1.4 ± 1.7 | 0.9 ± 1.1 |
| Flatulence (cm) | AG1 | 1.3 ± 1.5 | 1.3 ± 1.5 | 1.1 ± 1.2 | 1.1 ± 1.3 | 1.1 ± 1.5 |
|  | PL | 1.3 ± 1.9 | 1.3 ± 1.8 | 1.8 ± 2.3 | 1.5 ± 2.3 | 1.0 ± 1.6 |
| Hunger (cm)* | AG1 | 3.4 ± 2.2 | 3.5 ± 2.4 | 5.0 ± 2.1<sup>\$β</sup> | 5.4 ± 2.1<sup>\$β</sup> | 5.7 ± 1.8<sup>\$β</sup> |
| | PL | 4.4 ± 2.2 | 4.5 ± 2.0 | 5.2 ± 2.5 | 6.0 ± 2.0<sup>\$β</sup> | 6.1 ± 2.1<sup>\$β&</sup> |
| Appetite (cm) | AG1 | 3.6 ± 2.3 | 4.1 ± 2.3 | 5.0 ± 2.2<sup>\$β</sup> | 5.5 ± 2.0<sup>\$β</sup> | 5.6 ± 1.8<sup>\$</sup> |
| | PL | 4.1 ± 2.3 | 4.7 ± 2.2 | 5.4 ± 2.5<sup>\$</sup> | 6.1 ± 2.0<sup>\$β</sup> | 6.2 ± 2.0<sup>\$β</sup> |
| Fullness (cm) | AG1 | 3.4 ± 2.5 | 3.5 ± 2.5 | 3.2 ± 2.2 | 3.3 ± 2.3 | 3.2 ± 2.3 |
|  | PL | 2.8 ± 2.2 | 2.7 ± 1.8 | 2.6 ± 1.5 | 2.5 ± 1.6 | 2.4 ± 1.8 |
| Prospective food consumption (cm) | AG1 | 5.0 ± 2.7 | 5.1 ± 2.3 | 5.6 ± 1.9 | 5.9 ± 1.7<sup>\$</sup> | 6.2 ± 1.8<sup>\$β</sup> |
|  | PL | 5.7 ± 2.2 | 5.8 ± 2.2 | 5.7 ± 2.6 | 6.2 ± 2.3 | 6.8 ± 2.0 |
| Mood (cm) | AG1 | 6.8 ± 1.5 | 6.7 ± 1.4 | 6.7 ± 1.6 | 6.8 ± 1.5 | 7.0 ± 1.2 |
|  | PL | 7.3 ± 1.1 | 6.9 ± 1.5 | 6.6 ± 2.2 | 6.8 ± 2.2 | 7.0 ± 1.9 |
| Energy (cm) | AG1 | 5.7 ± 1.5 | 5.8 ± 1.5 | 5.9 ± 1.8 | 6.2 ± 1.8 | 6.3 ± 1.6 |
|  | PL | 5.9 ± 1.8 | 5.5 ± 1.7 | 5.5 ± 1.7 | 5.8 ± 1.8 | 6.0 ± 1.8 |
| Sleepiness (cm) | AG1 | 4.1 ± 1.8 | 3.8 ± 1.7 | 3.4 ± 1.9 | 3.3 ± 1.7 | 3.5 ± 2.0 |
|  | PL | 4.7 ± 2.2 | 4.4 ± 2.5 | 4.1 ± 2.0 | 4.0 ± 1.9 | 3.0 ± 1.6 |

Values are presented as mean ± SD for the total sample (n = 16). \* Indicates a significant difference between groups (p<0.05). <sup>\$</sup> Indicates a significant difference from 0 min (p<0.05). <sup>β</sup> Indicates a significant difference from 30 min (p<0.05). <sup>&</sup> Indicates a significant difference from 60 min (p<0.05).

#### 3.3.2 Diet and Safety

Assessment of total calories, protein, fat, or carbohydrates revealed no significant differences between groups (p>0.05) and were similar on both visits (Table 3). No differences in body weight or urine specific gravity were observed (Table 3). No significant difference was observed for systolic or diastolic blood pressure (Table 4). The study personnel observed two moderate adverse events from two separate subjects during the placebo condition of the trial (Table S4). One subject regurgitated at approximately 90 minutes following ingestion of the placebo, likely attributed to the investigational product, and one subject reported vasovagal responses at 0, 30, 90, and 240 minutes during the placebo condition, likely attributed to the blood draws.

**Table 3.**
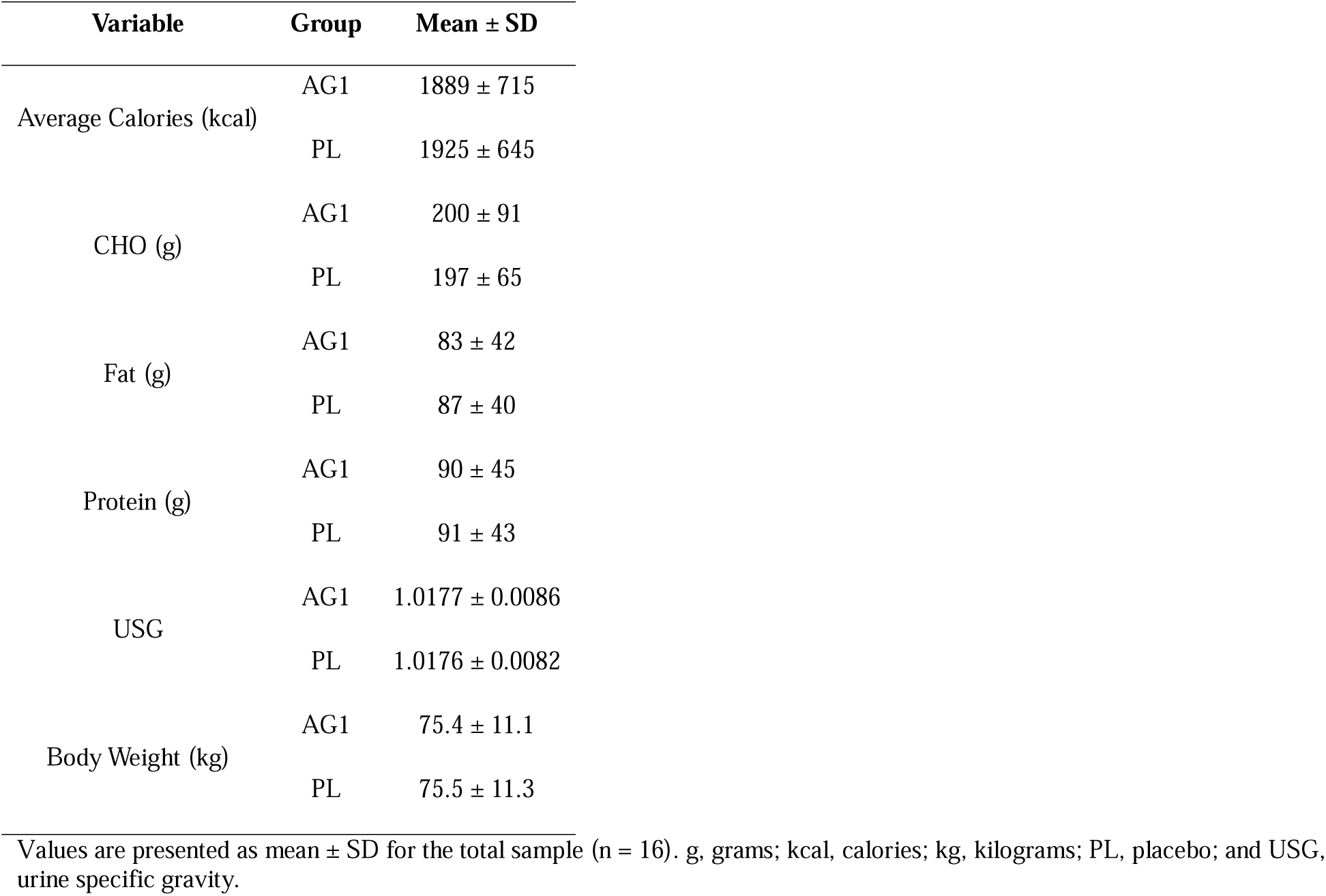
Dietary intake, urine specific gravity, and bodyweight at baseline.

**Table 4.** Blood pressure and heart rate responses over 8-hours post treatment administration.

| Variable | Group | 0 min | 60 min | 120 min | 180 min | 240 min | 360 min | 480 min |
| --- | --- | --- | --- | --- | --- | --- | --- | --- |
| SBP (mmHg) | AG1 | 117.3 ± 9.4 | 113.4 ± 10.2 | 117.1 ± 9.7 | 115.3 ± 8.9 | 115.0 ± 10.2 | 114.2 ± 12.5 | 113.9 ± 11.2 |
|  | PL | 116.9 ± 10.0 | 109.3 ± 10.7 | 114.9 ± 10.9 | 115.9 ± 8.4 | 112.8 ± 8.8 | 113.5 ± 11.1 | 114.6 ± 9.5 |
| DBP (mmHg) | AG1 | 71.8 ± 7.1 | 72.0 ± 9.4 | 69.1 ± 9.8 | 69.3 ± 10.1 | 70.0 ± 8.9 | 69.2 ± 11.7 | 68.8 ± 9.8 |
|  | PL | 71.8 ± 10.5 | 68.9 ± 8.8 | 69.8 ± 9.7 | 70.3 ± 9.7 | 66.9 ± 9.5 | 66.3 ± 9.5 | 69.2 ± 11.0 |
| HR (bpm) | AG1 | 63.9 ± 11.9 | 62.0 ± 10.0 | 60.3 ± 8.7 | 58.5 ± 8.8 | 62.5 ± 9.4 | 64.3 ± 8.9 | 65.6 ± 9.2 |
|  | PL | 63.6 ± 12.4 | 62.6 ± 11.6 | 62.4 ± 9.9 | 61.6 ± 12.5 | 60.4 ± 9.9 | 66.2 ± 12.5 | 63.7 ± 12.5 |

Values are presented as mean ± SD for the total sample (n = 16).BPM, beats per minute; DBP, diastolic blood pressure; min, minutes; mmHg, millimeters of mercury; PL, placebo; and SBP, systolic blood pressure.

## 4. Discussion

This randomized, double-blind, placebo-controlled crossover trial demonstrates that acute ingestion of AG1, which contains vitamins, minerals, and other nutrients, results in measurable systemic absorption of several vitamins, minerals, and phytochemicals. Compared to placebo, AG1 significantly elevated plasma concentrations following consumption and overall exposure (AUCL-LLL) for multiple nutrients, including folate, vitamin C, calcium, zinc, biotin, nicotinamide, and several B-vitamins, without any observed adverse events or tolerability concerns. These findings confirm that complex, multi-ingredient nutritional formulations can deliver physiologically relevant levels of bioactives to the bloodstream, satisfying the fundamental prerequisite for any systemic health benefit.

Absorption and bioavailability of nutrients may be influenced by nutrient-nutrient [17]and nutrient-matrix (e.g., delivery format, texture, etc.) interactions [18]. Vitamin C, for instance, is known to enhance non-heme iron absorption by reducing ferric iron to its more soluble ferrous form in the duodenum [21], while minerals like calcium and zinc can compete for absorption via shared transporters at the brush border membrane [22]. These synergistic and antagonistic interactions are well-documented, yet they become increasingly complex as the number of co-ingested nutrients grows. Prebiotics, probiotics, and plant-based fibers are components that may further modulate nutrient bioaccessibility by altering intestinal pH, enzyme activity, or microbiome composition [23,24]. These matrix effects underscore the importance of empirical data on nutrient pharmacokinetics within multi-ingredient formulations, as theoretical predictions based on single-nutrient data may not reflect real-world bioavailability.

Among the nutrients assessed in the present study, folate and vitamin C displayed the most consistent and robust pharmacokinetic profiles. Plasma concentrations rose significantly from baseline and remained elevated for several hours, resulting in substantial increases in both the peak serum concentrations and overall systemic exposure. These results align with findings from prior studies demonstrating efficient absorption and sustained circulation of liposomal vitamin C and non-liposomal formulations [15,16]. For calcium and zinc, peak plasma concentrations were observed later (between 30–60 min for calcium, between 60–240 min for zinc), with elevated levels persisting up to 240 minutes post-ingestion. Notably, levels of these minerals declined to baseline by 360 minutes contrasting the more sustained profiles seen with folate and vitamin C. Despite these temporal differences, both minerals exhibited significantly higher AUCs and peak concentrations following AG1 consumption compared to placebo. These distinct absorption profiles likely reflect nutrient-specific transport mechanisms such as passive diffusion for vitamin C, sodium-dependent co-transport for folate, and saturable ion channels for divalent cations like calcium and zinc, as well as potential competition from other nutrients or matrix components. However, in comparison to single ingredient pharmacokinetic data, the present study observed a higher C_max_ and shorter T_max_ than previous work administering the same mineral salts (e.g., calcium citrate, zinc citrate) [25] and various other different mineral salt forms of calcium (e.g., carbonate, pyruvate) and zinc (e.g., acetate, oxide) at doses substantially higher than contained in AG1 [26–29]Although differences in biochemical analysis methods limit direct comparison with previously reported pharmacokinetic data, the more soluble powder format of AG1 likely bypasses the digestive dissociation process, enhancing the bioaccessibility of its constituent nutrients compared to the tablets used in earlier studies [19]. Other factors, such as fasted versus fed states, can additionally alter pharmacokinetic parameters [30], underscoring the need for more pharmacokinetic research on nutrients and supplemental products.

The absorption kinetics of B vitamins varied widely. Thiamine (B1) showed the broadest temporal profile, with elevated levels from 30 to 360 minutes. In contrast, pyridoxine (B6) and riboflavin (B2) peaked only at 30 minutes, with no significant increases in AUC_0-480 min_ or C_max_. This discrepancy likely stems from biomarker selection: plasma pyridoxine and riboflavin are less sensitive to acute intake due to rapid conversion to active forms (e.g., pyridoxal 5’-phosphate) or preferential urinary excretion [31,32]. Nevertheless, AG1 produced significant elevations in thiamine, pyridoxine, and riboflavin while administering doses much lower than previously investigated in literature to date [33–37]. Future studies may consider urine biomarkers or metabolite profiling to potentially better capture short-term uptake of these vitamins [34]. Biotin and nicotinamide demonstrated intermediate absorption kinetics, with significantly elevated plasma levels and AUCs. As biotin is absorbed via both passive diffusion and the sodium-dependent multivitamin transporter (SMVT), potential competition with co-formulated pantothenic acid may influence its bioavailability [38]. However, this hypothesis warrants a more targeted subsequent investigation and was outside the scope of this study. Taken together, this study provides novel evidence that these b-vitamins have the capacity to significantly elevate blood nutrient levels at supplemental doses lower than previously reported and future work is needed to characterize nutrient pharmacokinetics at doses lower than typically consumed in commercially available single ingredient supplements.

The citrus bioflavonoid hesperidin, a phytochemical known for its antioxidant and vascular-modulating properties [39], is included in the AG1 formulation. Given the generally low bioaccessibility and variable absorption of flavonoid glycosides [40], hesperidin was assessed in this study on an exploratory basis. Hesperidin showed a transient but statistically significant elevation at 180 minutes post-AG1 ingestion, consistent with its presence in the product. Given the recognized challenges of flavonoid bioavailability, particularly for glycoside forms, these preliminary results are encouraging and warrant follow-up studies using additional conjugate-specific assays to better characterize hesperidin-derived bioactivity.

The trial was intentionally conducted in a fasted state, and participants were prompted to repeat the same dietary intake the day before testing in both conditions, to isolate AG1’s absorption profile from dietary confounders. Gastric emptying is typically accelerated under fasting conditions, and this may have contributed to the rapid appearance of certain nutrients in circulation [41]. However, the low enzymatic and bile activity during fasting may also impair digestion of fat-soluble or matrix-bound compounds [30,42]. Postprandial hepatic blood flow is also elevated, which may influence nutrient metabolism and first-pass clearance [43,44]. While the fasted-state design improves interpretability, it limits generalizability. Prior studies have shown that fed conditions can either suppress, enhance, or exert a minimal effect on absorption depending on nutrient or pharmaceutical form and food matrix composition [30,45–48]. Thus, future trials should evaluate AG1 in a fed state and compare its performance to single-nutrient reference formulations.

No adverse events were observed or reported for the AG1 treatment group, while two minor events were reported for the placebo treatment. Additionally, the results of the VAS in this trial did not reveal any significant adverse effects of AG1 on gastrointestinal symptoms (e.g., bloating and flatulence). Taken together, these data provide evidence that AG1, consumed acutely, is likely safe and well tolerated.

Important to note, this trial has several limitations. Traditional pharmacokinetic studies assess absorption, metabolism, distribution, and excretion whereas this study focused on acute absorption. However, this is appropriate for the assessment of MVM supplements given the intricate nuances of factors such as nutrient homeostasis, sex differences, and other biological factors. Furthermore, while the present study measured several key vitamins, minerals, and bioactives, full analysis of every nutrient in AG1 was outside the scope of this study. Additionally, while nutrient–nutrient antagonism was not formally evaluated, future studies should include co-analysis of inhibitors such as phytate and oxalate to characterize matrix effects more fully.

## 5. Conclusions

This study provides the first controlled pharmacokinetic evidence that the vitamins, minerals, and bioactives in a complex powdered MVM formulation (AG1) are absorbed into systemic circulation in physiologically meaningful amounts after a single dose for an acute period of time. Key micronutrients such as folate, vitamin C, calcium, and zinc displayed robust plasma appearance and favorable AUC profiles, indicating effective bioavailability. These findings fill a critical gap in the literature, which has historically focused on single-nutrient supplements, and offer a foundation for future research evaluating nutrient delivery, status, and clinical outcomes from complex multi-ingredient formulations.

## Data Availability

The dataset presented in this article are not readily available. Request to access the datasets should be directed to P.A.S.

