## Supplementary material for "Absorption Kinetics of Vitamins and Minerals from a Novel Nutritional Product in Healthy Adults: A Randomized, Double Blind, Placebo-Controlled Crossover Trial": Table S1

Table S1. Optimization details for B-vitamins, Biotin, and Hesperidin

|  | *Ion*  *mode* | *Precursor*  *Ion (m/z)* | *Product Ion (m/z)* | *Dwell time* | *Q1 Pre Bias (V)* | *CE* | *Q3 Pre Bias (V)* |
| --- | --- | --- | --- | --- | --- | --- | --- |
| *Thiamine* | *+* | *264.8* | *122.05* | *10* | *-30* | *-13* | *-22* |
| *pyridoxine* | *+* | *170.3* | *152.1* | *20* | *-12* | *-21* | *-24* |
| *Pyridoxine-d3* | *+* | *172.9* | *155.1* | *10* | *-21* | *-15* | *-29* |
| *Nicotinamide* | *+* | *123.1* | *80.0* | *10* | *-24* | *-22* | *-15* |
| *Nicotinamide-d4* | *+* | *126.9* | *84.0* | *10* | *-24* | *-22* | *-14* |
| *Riboflavin* | *+* | *377.1* | *243.0* | *30* | *-30* | *-25* | *-25* |
| *Riboflavin-d7* | *+* | *384.2* | *250.1* | *10* | *-30* | *-26* | *-27* |
| *Biotin* | *+* | *244.9* | *227.1* | *10* | *-13* | *-14* | *-25* |
| *Hesperidin* | *+* | *611.0* | *303.1* | *25* | *-28* | *-25* | *-21* |

m/z, mass to charge ratio; V, voltage
