## Supplementary material for "Absorption Kinetics of Vitamins and Minerals from a Novel Nutritional Product in Healthy Adults: A Randomized, Double Blind, Placebo-Controlled Crossover Trial": Table S2

Table S2. Clinical chemistry and hematological biomarkers at baseline.

| Variable | Mean ± SD |
| --- | --- |
| WBC (x10E3/uL) | 6.0 ± 2.5 |
| RBC (x10E6/uL) | 4.8 ± 0.5 |
| Hemoglobin (g/dL) | 14.0 ± 1.6 |
| Hematocrit (%) | 41.8 ± 4.0 |
| Glucose (mg/dL) | 90.8 ± 5.0 |
| BUN (mg/dL) | 13.6 ± 4.8 |
| Creatinine (mg/dL) | 1.0 ± 0.2 |
| BUN/Creatinine ratio | 98.5 ± 15.8 |
| eGFR (mL/min/1.73) | 14.7 ± 6.1 |
| Sodium (mmol/L) | 139.3 ± 1.3 |
| Potassium (mmol/L) | 4.4 ± 0.2 |
| Chloride (mmol/L) | 103.3 ± 2.0 |
| CO_2_ (mmol/L) | 23.2 ± 1.8 |
| Calcium (mg/dL) | 9.5 ± 0.4 |
| Total Protein (g/dL) | 7.0 ± 0.4 |
| Albumin (g/dL) | 4.4 ± 0.3 |
| Globulin (g/dL) | 2.5 ± 0.3 |
| A/G ratio (au) | 1.8 ± 0.2 |
| Bilirubin (mg/dL) | 0.5 ± 0.2 |
| Alkaline Phosphatase (IU/L) | 72.8 ± 22.7 |
| AST (IU/L) | 18.3 ± 6.3 |
| ALT (IU/L) | 20.6 ± 9.7 |
| Total Chol (mg/dL) | 173.2 ± 39.8 |
| Triglycerides (mg/dL) | 68.2 ± 25.4 |
| HDL (mg/dL) | 57.8 ± 10.3 |
| VLDL (mg/dL) | 13.3 ± 3.8 |
| LDL (mg/dL) | 102.1 ± 36.6 |
| LDL/HDL | 1.8 ± 0.7 |
| Total/HDL | 3.0 ± 0.7 |

Values are presented as mean ± SD for the total sample (n = 16). A/G ratio, albumin-to-globulin ratio; Albumin, serum albumin; ALT, alanine aminotransferase; Alkaline Phosphatase, alkaline phosphatase; AST, aspartate aminotransferase; BUN, blood urea nitrogen; BUN/Creatinine ratio, blood urea nitrogen to creatinine ratio; eGFR, estimated glomerular filtration rate; g/dL, grams per deciliter; HDL, high-density lipoprotein; IU/L, international units per liter; LDL, low-density lipoprotein; LDL/HDL, low-density lipoprotein to high-density lipoprotein ratio; mg/dL, milligrams per deciliter; mmol/L, millimoles per liter; RBC, red blood cell count; Total Chol, total cholesterol; Total/HDL, total cholesterol to high-density lipoprotein ratio; VLDL, very-low-density lipoprotein; WBC, white blood cell count; x10³/µL, thousands per microliter; x10⁶/µL, millions per microliter.
