## Supplementary material for "Absorption Kinetics of Vitamins and Minerals from a Novel Nutritional Product in Healthy Adults: A Randomized, Double Blind, Placebo-Controlled Crossover Trial": Table S3

Table S3. Pharmacokinetic results between conditions for all measured nutrients.

| **Variable** | **AG1** | **PL** | **P-value** |
| --- | --- | --- | --- |
| Folate | | | |
| AUC_0-480_ (ng/mL/min) | 8124 ± 1297 | 5489 ± 1723 | < 0.001 |
| C_max_ (ng/mL) | 20.8 ± 0.7 | 12.9 ± 3.9 | < 0.001 |
| T_max_ (min) | 37.5 ± 17.3 | 174 ± 111 | < 0.001 |
| Calcium | | | |
| AUC_0-480_ (mg/dL/min) | 4448 ± 116 | 4366 ± 146 | < 0.01 |
| C_max_ (mg/mL) | 9.51 ± 0.26 | 9.38 ± 0.3 | < 0.05 |
| T_max_ (min) | 129 ± 88.8 | 229 ± 167 | < 0.05 |
| Zinc | | | |
| AUC_0-480_ (μg/dL/min) | 41,140 ± 2,938 | 35,688 ± 3,353 | < 0.001 |
| C_max_ (μg/mL) | 112 ± 14.0 | 87.1 ± 8.8 | < 0.001 |
| T_max_ (min) | 150 ± 68.0 | 104 ± 88.5 | n.s. |
| Vitamin C | | | |
| AUC_0-480_ (mg/dL/min) | 598 ± 276 | 358 ± 206 | < 0.001 |
| C_max_ (mg/dL) | 1.55 ± 0.71 | 0.93 ± 0.49 | < 0.001 |
| T_max_ (min) | 146 ± 96 | 135 ± 144 | n.s. |
| Biotin | | | |
| AUC_0-480_ (ng/mL/min) | 404.4 ± 298.3 | 154 ± 225.1 | < 0.0001 |
| C_max_ (ng/mL) | 2.26 ± 1.928 | 0.57 ± 0.684 | 0.0036 |
| T_max_ (min) | 58 ± 32 | 139 ± 181 | n.s. |
| Nicotinamide | | | |
| AUC_0-480_ (ng/mL/min) | 6308.4 ± 7341.9 | 3613.4 ± 6157.2 | < 0.05 |
| C_max_ (ng/mL) | 81.6 ± 52.5 | 38.77 ± 39.7 | < 0.01 |
| T_max_ (min) | 53 ± 57 | 116 ± 65 | < 0.5 |
| Pyridoxine | | | |
| AUC_0-480_ (ng/mL/min) | 4.32 ± 4.1 | 2.19 ± 4.3 | n.s. |
| C_max_ (ng/mL) | 0.1 ± 0.07 | 0.07 ± 0.09 | n.s. |
| T_max_ (min) | 58 ± 84 | 66 ± 69 | n.s. |
| Riboflavin | | | |
| AUC_0-480_ (ng/mL/min) | 3271.4 ± 6319.3 | 1434.9 ± 5058.7 | < 0.001 |
| C_max_ (ng/mL) | 19.6 ± 25.7 | 15.2 ± 40.9 | n.s. |
| T_max_ (min) | 79 ± 83 | 206 ± 143 | < 0.05 |
| Thiamine | | | |
| AUC_0-480_ (ng/mL/min) | 3047.9 ± 2865.4 | 47.9 ± 116.8 | < 0.001 |
| C_max_ (ng/mL) | 17.35 ± 13.03 | 2.32 ± 2.49 | < 0.0001 |
| T_max_ (min) | 84 ± 49 | 174 ± 160 | < 0.05 |
| Hesperidin | | | |
| AUC_0-480_ (ng/mL/min) | 27.99 ± 70 | 5.32 ± 10.65 | n.s. |
| C_max_ (ng/mL) | 0.28 ± 0.75 | 0.07 ± 0.7 | n.s. |
| T_max_ (min) | 129 ± 144 | 122 ± 151 | n.s. |

Values are presented as mean ± SD for the total sample (n = 16). AUC_0-480_, area under the curve 0-480 minutes; C_max_, maximum concentration; min, minutes; n.s., not significant; PL, placebo; and T_max_, time to maximum concentration.
