## Supplementary material for "Absorption Kinetics of Vitamins and Minerals from a Novel Nutritional Product in Healthy Adults: A Randomized, Double Blind, Placebo-Controlled Crossover Trial": Table S4

Table S4. Adverse events results.

|  | PL  (n=16) | AG1 (n=16) |
| --- | --- | --- |
| **Severity** |  |  |
| Mild | 0 | 0 |
| Moderate | 2 | 0 |
| Severe | 0 | 0 |
| **Relationship to Study Treatment** |  |  |
| Unlikely | 0 | 0 |
| Possible | 0 | 0 |
| Probable | 1 | 0 |
| **Relationship to Testing Procedures** |  |  |
| Unlikely | 0 | 0 |
| Possible | 0 | 0 |
| Probable | 1 | 0 |
| N/A | 0 | 0 |
| **Body System and AEs** |  |  |
| Gastrointestinal | 0 | 0 |
| Regurgitation | 1 | 0 |
| Cardiovascular | 0 | 0 |
| Vasovagal | 1 | 0 |
| Total Number of Adverse Events Experienced During Study | 2 | 0 |
| Total Number of Subjects Experiencing Adverse Events: n (%) | 2/17 (~11.8%) | 0/16 (0%) |

Values are presented as the total number of participants for the total sample (n = 16).
